# Multivariate whole brain neurodegenerative-cognitive-clinical severity mapping in the Alzheimer’s disease continuum using explainable AI

**DOI:** 10.1101/2025.07.09.25331194

**Authors:** Taslim Murad, Hui-Yuan Miao, Deepa S. Thakuri, Gauri Darekar, Alzheimer’s Disease Neuroimaging Initiative, Ganesh B. Chand

## Abstract

Neurodegeneration and cognitive impairment are commonly reported in Alzheimer’s disease (AD); however, their multivariate links are not well understood. To map the multivariate relationships between whole brain neurodegenerative (WBN) markers, global cognition, and clinical severity in the AD continuum, we developed the explainable artificial intelligence (AI) methods, validated on semi-simulated data, and applied the outperforming method systematically to large-scale experimental data (N=1,756). The outperforming explainable AI method showed robust performance in predicting cognition from regional WBN markers and identified the ground-truth simulated dominant brain regions contributing to cognition. This method also showed excellent performance on experimental data and identified several prominent WBN regions hierarchically and simultaneously associated with cognitive declines across the AD continuum. These multivariate regional features also correlated with clinical severity, suggesting their clinical relevance. Overall, this study innovatively mapped the multivariate regional WBN-cognitive-clinical severity relationships in the AD continuum, thereby significantly advancing AD-relevant neurobiological pathways.

## Introduction

Alzheimer’s disease (AD) is a multifactorial neurodegenerative disorder and the most common cause of dementia [1]. Its prevalence is over 55 million people worldwide [2] and is becoming an alarming global health concern [3]. It causes cognitive declines, behavioral alterations, disorientation, and memory loss in affected individuals, leading to reduced quality of life [4] and accelerated progression to death [5]. The rising personal and financial burdens associated with AD require investigations across the AD continuum, thereby fully understanding disease pathways and potentially providing effective pre-clinical or clinical diagnoses and treatments to prevent disease progression. The complex pathogenesis of AD may stem from a combination of various factors, including amyloid-beta (Aβ), tau, alpha-synuclein, genetic, epigenetic, and environmental, among others, making it difficult to fully comprehend. AD may manifest cognitive impairment due to loss of synaptic function [6] and atrophy in the limbic system, neocortex, and basal forebrain [7, 8]. The Aβ plaques and tau protein tangles in the brain are also key for cognitive declines in AD, causing brain cell death by disrupting cell communication and triggering inflammatory responses [9, 10]. All these factors, among others, may aggregate to neurodegeneration processes. Hence, the neurodegeneration or neurostructural atrophy is the most proximate substrate of cognitive impairment in AD and is essential for unraveling the complexities of AD etiology and neurobiological pathways.

Capturing spatial mapping of neurodegeneration markers along the AD continuum using in vivo imaging biomarkers is crucial to identify neurobiological pathways and aid in disease diagnosis and prognosis. A key advantage of brain imaging techniques is their superior spatiotemporal sensitivity to brain structural atrophies compared to fluid biomarkers [11]. Among imaging biomarkers, structural magnetic resonance imaging (sMRI) is promising for quantifying brain spatial morphometry, as it assesses atrophy in gray matter, white matter, and cerebrospinal fluid (CSF)/ventricular regions, thereby offering a broader perspective on disease progression [12, 13], unlike its other variant, diffusion tensor imaging (DTI), which is sensitive to microstructural properties of white matter only [14]. Similarly, positron emission tomography (PET) molecular imaging techniques can measure Aβ-, tau-, or other protein-specific regional bindings associated with AD. However, in neurodegeneration research, the quantification of these proteins from all cortical and subcortical regions is still challenging due to partial volume spill in/out effects and off-target bindings, and these may lead to inaccuracies in assessing their true concentration and distribution within the diverse brain regions [15]. Critically, PET molecular imaging techniques are highly sensitive only to one tracer-specific protein (e.g., Aβ, tau, alpha-synuclein, or others), thereby ignoring other multifactorial aggregated contributions to AD and associated cognitive declines. The sMRI is a current state-of-the-art measurement that can provide aggregated effects of multifactorial bases of AD, providing holistic neurodegeneration spatial markers and their associations with cognitive declines in AD. Therefore, herein we employed sMRI and global cognition to investigate multivariate whole brain neurodegenerative (WBN)-cognition mappings in the AD continuum. Global cognition was assessed using the Mini-Mental State Examination (MMSE) [16], a 30-point questionnaire that evaluates multiple cognitive domains, including orientation, registration, attention and calculation, recall, and language. Lower MMSE scores indicate greater cognitive impairment.

The integration of advanced artificial intelligence (AI) algorithms and neuroimaging technologies has enormous potential for understanding disease mechanisms [17-21]. Several prior studies [22, 23] have successfully applied machine learning (ML)/ deep learning (DL) models to detect AD through classification or regression tasks; however, these approaches often do not investigate how specific brain regions contribute to the underlying aspect under study, such as cognitive decline, within the AD continuum. Some interpretable approaches [24-29] have attempted to address this gap using correlation analyses, mass-univariate methods, or linear mixed-effects models. However, these techniques typically assess the association between individual brain regions and cognitive scores in isolation, which limits their ability to capture the multivariate interactions across multiple brain regions that may jointly influence cognitive performance. Although functional neuroimaging studies (functional MRI, PET, local field potential, E/MEG and others) have widely shown that multiple brain regions are collectively involved in cognitive processes [30-40], implying the critical need for the development of new multivariate methodologies that can fully integrate neurodegeneration information from multiple regions simultaneously and identify the underlying neurodegeneration-cognitive deficit progression pathways. A prior data-driven explainable AI technique [41] has presented proof-of-concept results for capturing such multivariate relationships by combining a deep learning (DL) model with Shapley additive explanations (SHAP) [42] feature importance strategy on limited brain regions (Braak-stages; n=22) using a small and less diverse dataset (n=33) [43]. Although promising, this method remains to be fully validated on the whole brain regional features extracted from large neuroimaging samples belonging to diverse cohorts for its broader generalizability. Additionally, this method remains to be extended to investigate clinical severity aspects of AD, as while clinical severity trajectories are observed in AD, the mechanisms underlying their onset and progression pathways remain poorly understood.

In this study, the predictive performances of several AI models were systematically investigated and compared for predicting global cognition using regional neurodegeneration markers simultaneously from WBN spatial locations (n=145) obtained from large and diverse sMRI samples (N=1,756). The optimal model was further integrated with SHAP, to form DL-SHAP method, for identifying holistic multivariate regional neurodegeneration predictors for global cognition in the AD continuum. These individualized multivariate regional predictors were then correlated with the clinical severity metrics in patients and controls. We hypothesize that our explainable AI approach will identify neurodegeneration-related dominant regional hubs or network regions that are critical during the global cognitive impairment process, and those hubs will associate with clinical or disease severity measurements across the AD continuum.

## Materials and Methods Experimental data

The baseline T1-weighted sMRI data from the Alzheimer’s Disease Neuroimaging Initiative (ADNI)-2 [44] and Knight Alzheimer Disease Research Center (Knight-ADRC) [45] cohorts were employed in this study as an experimental data. We considered the participants with available baseline MMSE scores. The ADNI dataset has 668 participants (308 females; age: 55.1-91.5 years old, mean = 72.8 years old, standard deviation = 7.3 years old), where 187 are cognitively normal (CN), 6 are mild cognitively impaired (MCI), 173 are early MCI (EMCI), 152 are late MCI (LMCI), and 150 AD subjects. The Knight-ADRC data has 1,088 participants (605 females; age: 45.2-91.4 years old, mean = 70.4 years old, standard deviation = 8.5 years old) categorized into 920 CN and 168 MCI/AD subjects. Note that Knight-ADRC has only two diagnostic categories, with all the patients (LMCI/EMCI/MCI/AD) in the AD group. Since both data cohorts encompass participants across different age ranges, their integration facilitates the development of a more generalized model that accounts for age-related variability. The detailed summary of the experimental data is given in **Table 1**.

**Table 1:** Experimental data. Distribution of participants by diagnosis and sex in the ADNI and Knight-ADRC data cohorts. Descriptive statistics for age and MMSE scores are provided for each cohort, including minimum (Min), maximum (Max), mean (Mean), and standard deviation (Std).

| Data Cohort | Diagnosis |  |  |  |  | Sex |  | Age (years) |  |  |  | MMSE Score |  |  |  |
| --- | --- | --- | --- | --- | --- | --- | --- | --- | --- | --- | --- | --- | --- | --- | --- |
|  | CN | EMCI | LMCI | MCI | AD | Male | Female | Min | Max | Mean | Std | Min | Max | Mean | Std |
| ADNI | 187 | 173 | 152 | 6 | 150 | 360 | 308 | 55.1 | 91.5 | 72.8 | 7.3 | 19 | 30 | 27.2 | 2.8 |
| Knight ADRC | 920 | 168 |  |  |  | 483 | 605 | 45.2 | 91.4 | 70.4 | 8.5 | 15 | 30 | 28.5 | 2.1 |

Moreover, to process the data, each sMRI was segmented into 145 anatomical regions of interest (ROIs), encompassing the entire brain’s gray matter, white matter, and cerebrospinal fluid (CSF)/ventricular regions. This segmentation was performed using the Multi-atlas region segmentation using Ensembles of registration algorithms and parameters and the Locally Optimal Atlas Selection (MUSE) method [46]. As a multi-atlas tool, MUSE has been shown to outperform traditional segmentation methods, particularly in subcortical regions and in diseased populations [47]. Extracting a large number of ROIs allows for a more fine-grained representation of the brain’s structural subdivisions, enabling more nuanced insights into WBN-cognition mappings. Additionally, neurodegenerative biomarkers were extracted from the ROIs and harmonized using an existing regression-based harmonization technique [21, 48, 49] to adjust for site, cohort, age, sex, and total intracranial volume for each participant. The harmonized data were then normalized by dividing each ROI volume by the total volume across all ROIs for that participant, thereby accounting for individual head size differences and making the data more suitable for AI-based predictive modeling. The resulting harmonized and normalized ROI features were used as input to AI models for predicting global cognitive (MMSE). Furthermore, clinical severity of MCI/AD was assessed using the Clinical Dementia Rating–Sum of Boxes (CDR-SB) scores [50, 51], recorded at the time point closest to each participant’s sMRI acquisition in both the ADNI and Knight ADRC cohorts.

### Semi-simulated data

We formulated semi-simulated data to validate the ability our DL-SHAP method for capturing the hierarchical multivariate WBN-cognition mappings. The objective of this simulation analysis was to construct arbitrary multivariate associations between brain neurodegenerative markers and the cognitive measure (MMSE), and to evaluate whether our method could accurately recover these simulated patterns. The semi-simulated data were generated using sMRIs of CN participants (n = 1,107) from the experimental dataset. Using only CN data allows for the creation of a clean baseline, where any deviations introduced through synthetic perturbations can be clearly interpreted as disease-like signals. Further details of the simulation setup are provided in supplementary section **S1**.

### Global cognition prediction with ML/AI models

We employed three conventional machine learning (ML) models and a deep learning (DL) model to perform global cognition (MMSE) predictions. Then the outperforming model was integrated with feature importance techniques, particularly SHAP [42] and others mentioned below, to capture the hierarchy of multivariate WBN-cognition mappings.

The ML models used here were Lasso regression (LR) [52, 53], Ridge regression (RR) [54, 55], and support vector regression (SVR) [19]. These models were selected because of their demonstrated efficacy in handling high-dimensional datasets. For each ML model, its hyperparameters were optimized following the grid search approach with negative mean absolute error scoring metric. For LR and RR models, the optimal alpha parameter was chosen from a range of 0.15-0.35 and 150-250 with an increment of 0.5 and 10, respectively. This parameter controls the strength of regularization applied to the model to prevent overfitting. For the SVR model, the Radial Basis Function (RBF) kernel was utilized, with hyperparameters gamma, epsilon, and C optimized over the following ranges: gamma from -12 to -2 (with an increment of 1), epsilon from -7 to 1 (with an increment of 1), and C from -1 to 4 (with an increment of 1). These hyperparameters work together to balance the model’s complexity, generalization ability, and sensitivity to training data.

Moreover, considering the superior capability of DL models in capturing complex non-linear relationships as compared to the conventional ML models, we tested MMSE prediction using a basic deep neural network (DNN)-based DL model. Given our data constraint (relatively not very large sample size for DL models), this model is the best fit as it provides promising generalized results while minimizing the risk of overfitting due to its architectural simplicity. Our DL model consists of four hidden neural network (NN) blocks and one output NN block. Each inner NN block has a dense layer, with rectified linear unit (ReLU) activation followed by a batch normalization and a dropout. Batch normalization alleviates the model’s performance by addressing the challenges like unstable activations, slow convergence, overfitting, and gradient problems. Dropout acts as a regularization to reduce the risk of overfitting by randomly deactivating a fraction of artificial neurons in each iteration. The architecture featured a decreasing number of units in the inner layers, specifically 40, 30, 20, and 10 units, respectively. The output NN block is a single dense unit with linear activation to predict the cognitive (MMSE) scores. The random normal initialization from the Keras library was employed to initialize the weights of each dense layer for introducing randomness and preventing the model from getting stuck in a local minimum during training. We also implemented an early stopping callback that keeps track of the validation loss function during training, halts it when it stops getting better, and restores the model’s best weights. The DL model was created using the Python’s TensorFlow and Keras libraries. To determine the optimal hyperparameters for the model, we performed hyperparameters tunning and its details are presented in supplementary section **S2**.

### DL-SHAP

The DL model portrayed optimal performance in predicting the MMSE scores as compared to the conventional ML models, therefore this model was integrated with SHAP, formed DL-SHAP, to capture the hierarchy of multivariate associations between WBN and cognition scores. SHAP is a popular widely used feature importance strategy that accounts for interactions among input features and is generally more stable and consistent than the other famous feature importance techniques, like local interpretable model-agnostic explanations (LIME) [56, 57] and layer-wise relevance propagation (LRP) [58], therefore we employed SHAP in our analysis. The detailed comparison of SHAP with other feature importance techniques (LIME and LRP) are provided in supplementary section **S3**. SHAP is a state-of-the-art approach for the interpretability and explainability of complex predictive models, providing a global perspective by revealing the overall significance of each input feature towards the prediction, hence explaining the black box models like DL models. It satisfies all three essential properties concerning interpretability, explainability, and accuracy of the black box AI model, and those properties are local accuracy, missingness, and consistency. Local accuracy ensures that the SHAP explanation model faithfully reproduces the original model’s prediction for a specific instance by guaranteeing that the sum of the SHAP values across all input features equals the output of the original model. When all features are set to their baseline (reference) values, the explanation model’s prediction matches the model’s expected prediction under the same condition. Missingness guarantees that the absence of certain features does not affect the interpretation, while consistency ensures that feature importance remains unchanged unless a feature’s contribution alters. SHAP quantifies the significance of an input feature for a model’s output through its SHAP value, which is derived from the Shapley value in cooperative game theory. Through Shapley value players in a game are assigned fair values based on their contribution to the game’s output. The SHAP value for an input feature *i* is computed through the given formula [41],

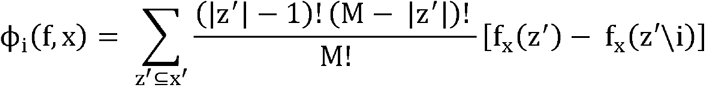

where *f* denotes the trained model, x = [x_1_, x_2,_ …, x_M_] represents the input features, and x, refers to simplified binary input vectors indicating which features are included (1) or excluded (0). Each subset z, of x, reflects a specific combination of present or absent features, and the term *f*_x_(z,) is the expected model output when only the features in z,are known. The difference f_x_(z,) -f_x_(z^,\^i) captures the marginal effect of including ensures fair contribution from each feature *i*, and the weighting factor 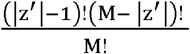 subset, derived from cooperative game theory principles. To illustrate, if we consider an input with three features A, B, and C, then all possible subsets of these features are represented by binary vectors such as [1, 0, 1] (including A and C only), and SHAP evaluates how the presence or absence of each feature affects the model’s prediction. For each feature, these contributions are aggregated across all possible subsets and weighted accordingly, providing a comprehensive measure of the feature’s importance while accounting for interactions with other features. The final SHAP score for a feature is computed by averaging the absolute SHAP values across all individuals in the dataset, and a higher average absolute value reflects greater influence of that feature on the model’s prediction. Thus, SHAP provides both local (sample-specific) and global (across all samples) interpretability, making it well-suited to identify important features and multivariate associations in models predicting cognitive outcomes like MMSE scores. The workflow of our proposed DL-SHAP method is shown in **Figure 1**. It performs global cognition (MMSE) prediction using the DL model based on the regional imaging features as input and computes the SHAP values of these input features to identify their hierarchical contribution towards the global cognition prediction.

**Figure 1:**
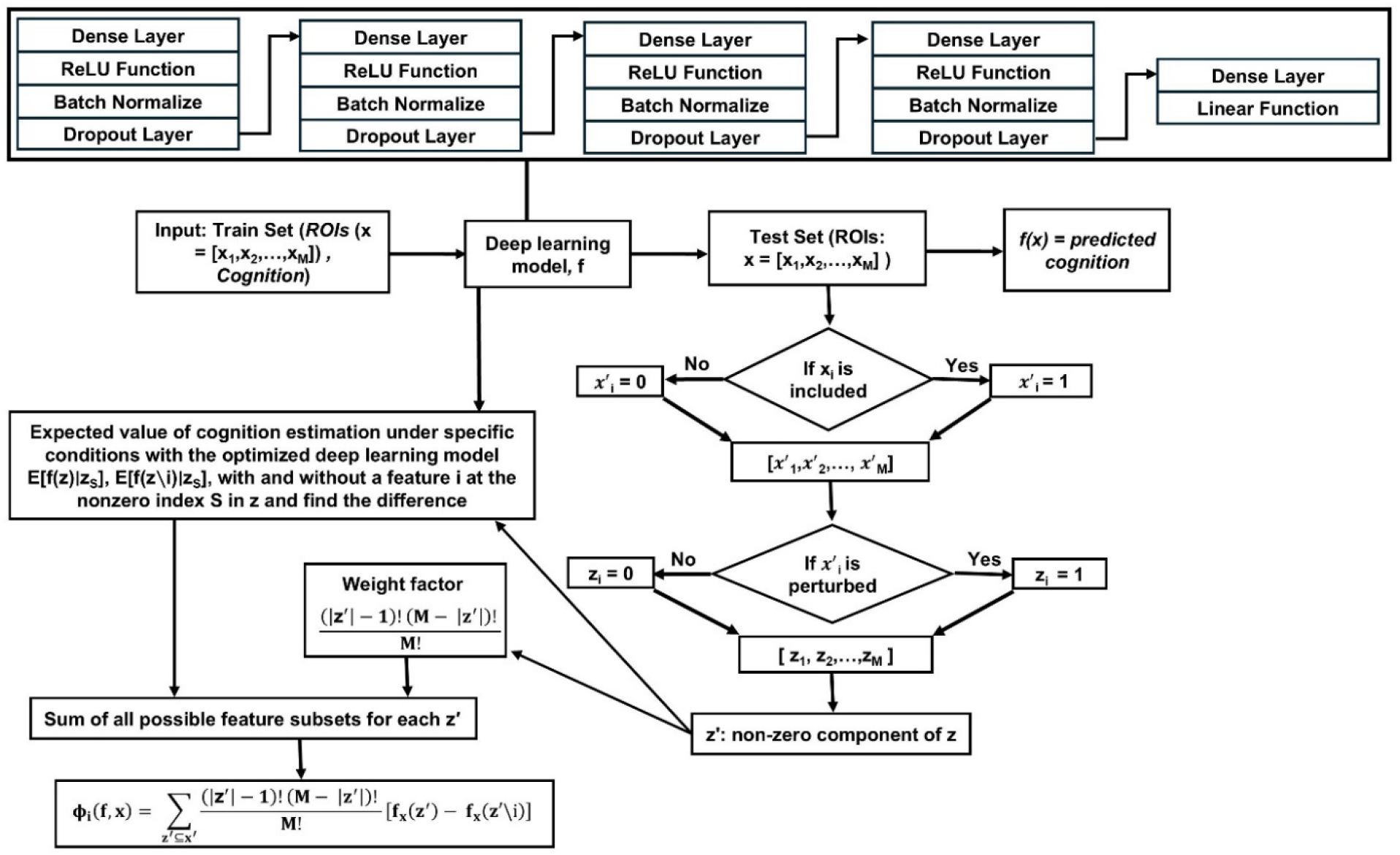
Schematic of the proposed DL-SHAP method.

To train and evaluate our models, we followed a 10-fold cross validation (CV) [59] procedure. In this the data was randomly partitioned into 10 folds, and in each fold, 90% of the data was used for training and 10% for testing, while ensuring no overlap between training and test datasets. For the DL model, the train data in every fold was further divided into 95% train dataset and 5% validation dataset. The validation set was used to monitor training (e.g., for early stopping) before evaluating the model on the test set. Note that, in each iteration of CV, there were no overlapping samples among train, test, and validation datasets. Furthermore, to ensure reproducibility and minimize noise, each model was executed five times, where each run consists of 10-fold CV training-testing process. Our reported results for each model are based on the independent test set and represent the mean performance across five separate runs. Additionally, to evaluate the model performance, we computed the Spearman correlation between the actual and predicted MMSE scores on the test set, accompanied with the *p*-value to validate the significance of the correlation, using Spearman rank-order correlation coefficient method [60]. This is a non-parametric approach which does not make assumptions about the underlying data distribution. We also reported the averaged mean absolute error (MAE) loss on the test set for each model. Moreover, all the figures based on brain slices are generated by overlaying the results on the standard MRI template (in MNI space), following an axial orientation for the visualization. For the experimental data, the brain figures are portrayed for CN, MCI and AD groups, where CN has only cognitively normal participants, AD consists of participants belonging to AD diagnosis category, and MCI is based on all the individuals having EMCI, LMCI or MCI diagnosis. Furthermore, the neurodegeneration differences between the CN and available patient groups in both cohorts were analyzed using the Cohen’s d effect size metrics [61]. Individualized DL-SHAP regional values were used to compute Cohen’s d. Only ROIs with p-values less than 0.05 were considered statistically significant. The Cohen’s d-based analysis aims to illustrate how brain alterations in terms of multivariate regional associations with cognition evolve as the disease progresses.

### Clinical severity under the DL-SHAP framework

We investigated the clinical severity of participants in AD continuum using the CDR-SB score. This score assesses the clinical or disease severity based on three cognitive and three functional domains (memory, orientation, judgement and problem solving, community affairs, home and hobbies, and personal care) [62] and its value spans from 0-18. A higher CDR-SB score indicates a greater clinical or disease severity. In our analysis, the Spearman’s correlation between CDR-SB scores and the individualized DL-SHAP regional features was explored to highlight the relevance of DL-SHAP identified features in determining the clinical severity of MCI/AD.

## Results

### Semi-simulated data results

The performances of the conventional ML models and the DL model for MMSE prediction using the semi-simulated data are given in **Table 2**. We can observe that the DL model outperformed the ML models by achieving a Spearman’s correlation of 0.940 between the actual and predicted MMSE scores **(Figure 2(a))**, whereas the LR, RR, and SVR models obtained a correlation of 0.894, 0.883, and 0.879, respectively. All correlations were statistically significant, with p-values less than 0.05. Additionally, the DL model also obtained the minimum test loss (MAE) of 0.872. Moreover, the DL-SHAP method was able to identify all the ground-truth perturbed brain regions **(Figure 2(b))** as the most significant brain regions for global cognition prediction based on their SHAP values, as demonstrated in **(Figure 2(c))**. Note that, **Figure 2(b)** highlights only the regions that were specifically perturbed to generate the semi-simulated data. In contrast, **Figure 2(c)** presents SHAP values, which were continuous and captured a hierarchy of feature importance. As a result, the perturbed regions in **Figure 2(c)** are visualized with varying color intensities to reflect their relative importance. Despite these variations, all highlighted regions were among the top 10 most significant contributors to global cognition prediction. This indicates that the DL-SHAP method consistently identified the perturbed regions as highly influential, even though their individual contributions to the prediction may vary.

**Table 2:**
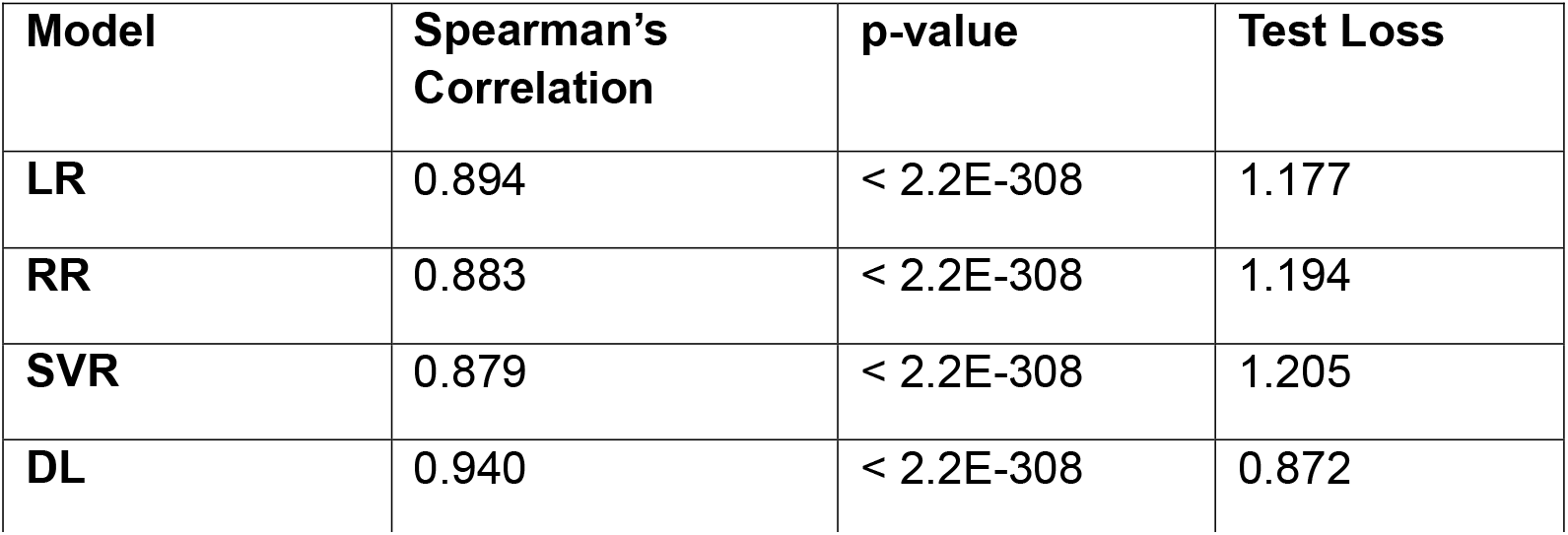
Semi-simulated dataset. The performance of Lasso regression (LR), Ridge regression (RR), support vector regression (SVR), and DL model for global cognition (MMSE) prediction.

| Model | Spearman's Correlation | p-value | Test Loss |
| --- | --- | --- | --- |
| LR | 0.894 | < 2.2E-308 | 1.177 |
| RR | 0.883 | < 2.2E-308 | 1.194 |
| SVR | 0.879 | < 2.2E-308 | 1.205 |
| DL | 0.940 | < 2.2E-308 | 0.872 |

**Figure 2:**
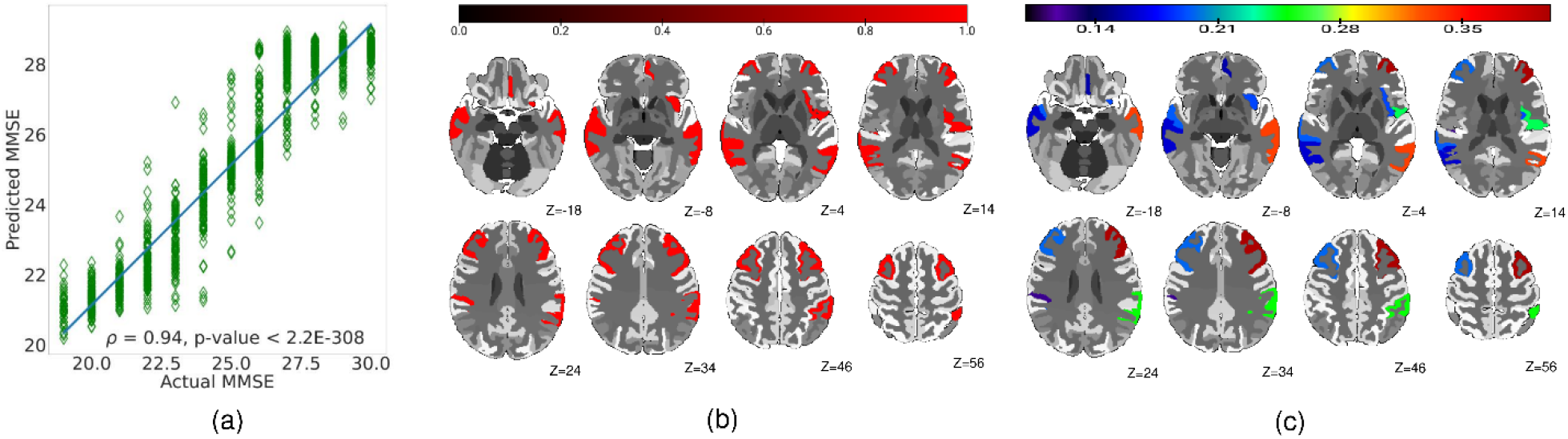
DL-SHAP on semi-simulated data. **(a)** Spearman’s correlation between the predicted and simulated MMSE scores. **(b)** The top 10 regions chosen for perturbation are shown in red. **(c)** Ground-truth simulated regions (averaged over all participants) identified as significant brain regions using DL-SHAP. The color bar represents SHAP values of ROIs, and z shows the brain slice position.

### Experimental data results

The experimental results were obtained by first training the models on the full experimental dataset (n=1,756) and then separating out the test samples from ADNI, Knight-ADRC, and the combined (ADNI + Knight-ADRC) data to evaluate and report the respective performance metrics. **Table 3** summarizes the performance of the ML models and the DL model for MMSE prediction on the ADNI, Knight-ADRC, and combined test data, respectively. On the combined test data, the DL model achieved a superior Spearman’s correlation of 0.935 between the actual and predicted MMSE scores, outperforming the LR, RR, and SVR models, which obtained correlations of 0.492, 0.529, and 0.585, respectively. All correlations were accompanied by extremely small p-values, indicating high statistical significance. Furthermore, the DL model yielded the lowest test loss (MAE = 0.675), suggesting better generalization to unseen data compared to the ML models. When evaluated on individual cohorts, the DL model continued to demonstrate strong performance. It achieved a correlation of 0.981 on the ADNI test data (**Figure 3(a)**) and 0.892 on the Knight-ADRC test data (**Figure 3(b)**). In contrast, the LR, RR, and SVR models produced correlations of 0.583, 0.581, and 0.618, respectively, on the ADNI set, and 0.351, 0.372, and 0.373, respectively, on the Knight-ADRC set. Note that because the models were trained on the full experimental dataset, the test loss was reported for the combined test set only.

**Table 3:** Experimental dataset. The performance of Lasso regression (LR), Ridge regression (RR), support vector regression (SVR), and deep learning (DL) models for predicting global cognition (MMSE).

| Model | Data Cohort | Spearman's Correlation | p-value | Test Loss |
| --- | --- | --- | --- | --- |
| LR | ADNI | 0.583 | 3.79E-62 | 1.655 |
|  | Knight-ADRC | 0.351 | 4.64E-33 |  |
|  | Combined | 0.492 | 5.81E-108 |  |
| RR | ADNI | 0.581 | 1.16E-61 | 1.631 |
|  | Knight-ADRC | 0.372 | 3.38E-37 |  |
|  | Combined | 0.529 | 2.13E-127 |  |
| SVR | ADNI | 0.618 | 9.62E-72 | 1.518 |
|  | Knight-ADRC | 0.373 | 2.39E-37 |  |
|  | Combined | 0.585 | 1.99E-162 |  |
| DL | ADNI | 0.981 | <2.20E-308 | 0.675 |
|  | Knight-ADRC | 0.892 | <2.20E-308 |  |
|  | Combined | 0.935 | <2.20E-308 |  |

**Figure 3:**
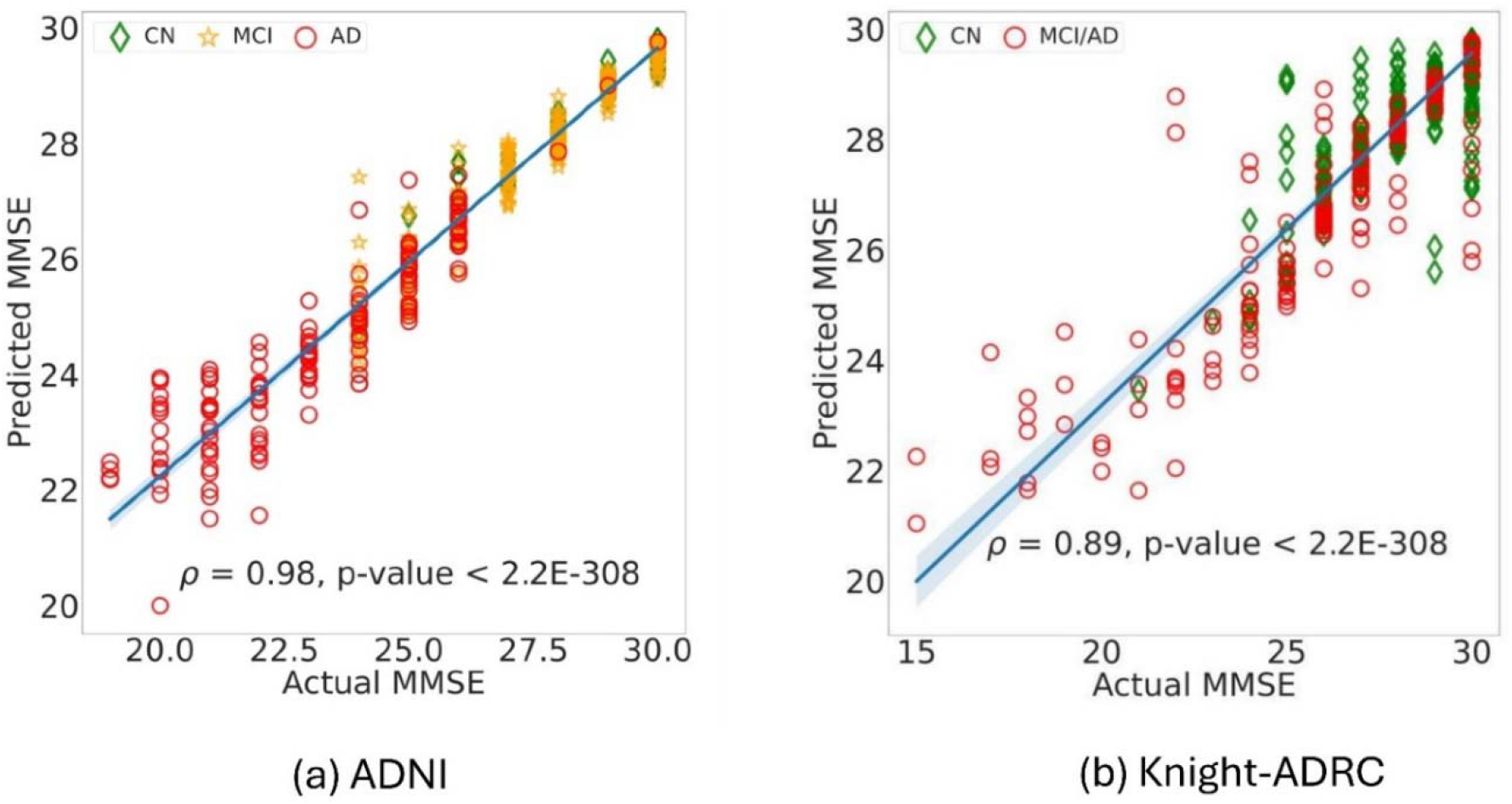
Spearman’s correlation between the actual and predicted MMSE scores using the DL model on **(a)** ADNI data and **(b)** Knight-ADRC data.

Moreover, for each data cohort, DL-SHAP identified the significant brain regions towards MMSE prediction in control and patient groups. For instance, **Figure 4(a)** and **Figure 5(a)** illustrate the dominant brain regions for predicting global cognition in the CN group of ADNI and Knight-ADRC data, respectively. In the ADNI CN group, the most significant regions included the left middle temporal gyrus, left inferior lateral ventricle, left hippocampus, right cerebellum white matter, and left occipital fusiform gyrus. Similarly, in the Knight-ADRC CN group, the dominant regions were the left middle temporal gyrus, right cerebellum white matter, left cerebellum white matter, left inferior lateral ventricle, and left occipital fusiform gyrus. Additionally, the dominant brain regions in the MCI group of ADNI were the left middle temporal gyrus, left inferior lateral ventricle, right cerebellum white matter, left cerebellum white matter, and left occipital fusiform gyrus, as illustrated in **Figure 4(b)**. The major brain regions in AD category of ADNI were the left middle temporal gyrus, left inferior lateral ventricle, left hippocampus, right cerebellum white matter, and left cerebellum white matter, as given in **Figure 4(c)**, while in MCI/AD group of Knight-ADRC were the left middle temporal gyrus, left inferior lateral ventricle, left hippocampus, right cerebellum white matter, and left cerebellum white matter, as given in **Figure 5(b)**.

**Figure 4:**
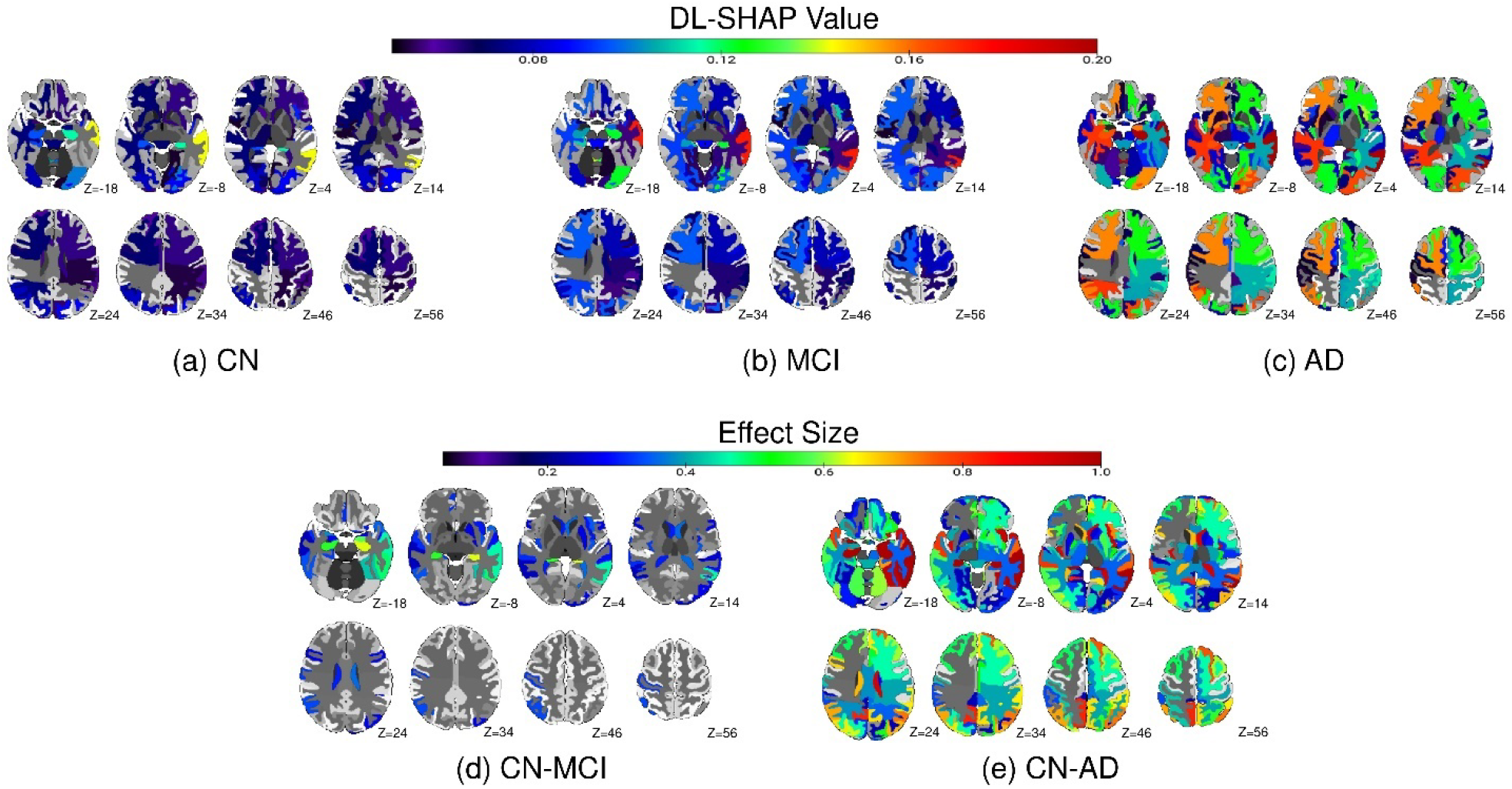
DL-SHAP identified significant brain regions for predicting global cognition (MMSE) using the **ADNI** experimental data across the **(a)** CN, **(b)** MCI, and **(c)** AD groups. The results are averaged over all participants. The Cohen’s d effect size differences in regional DL-SHAP values are shown between groups: **(d)** CN and MCI and **(e)** CN and AD. z shows the brain slice position.

**Figure 5:**
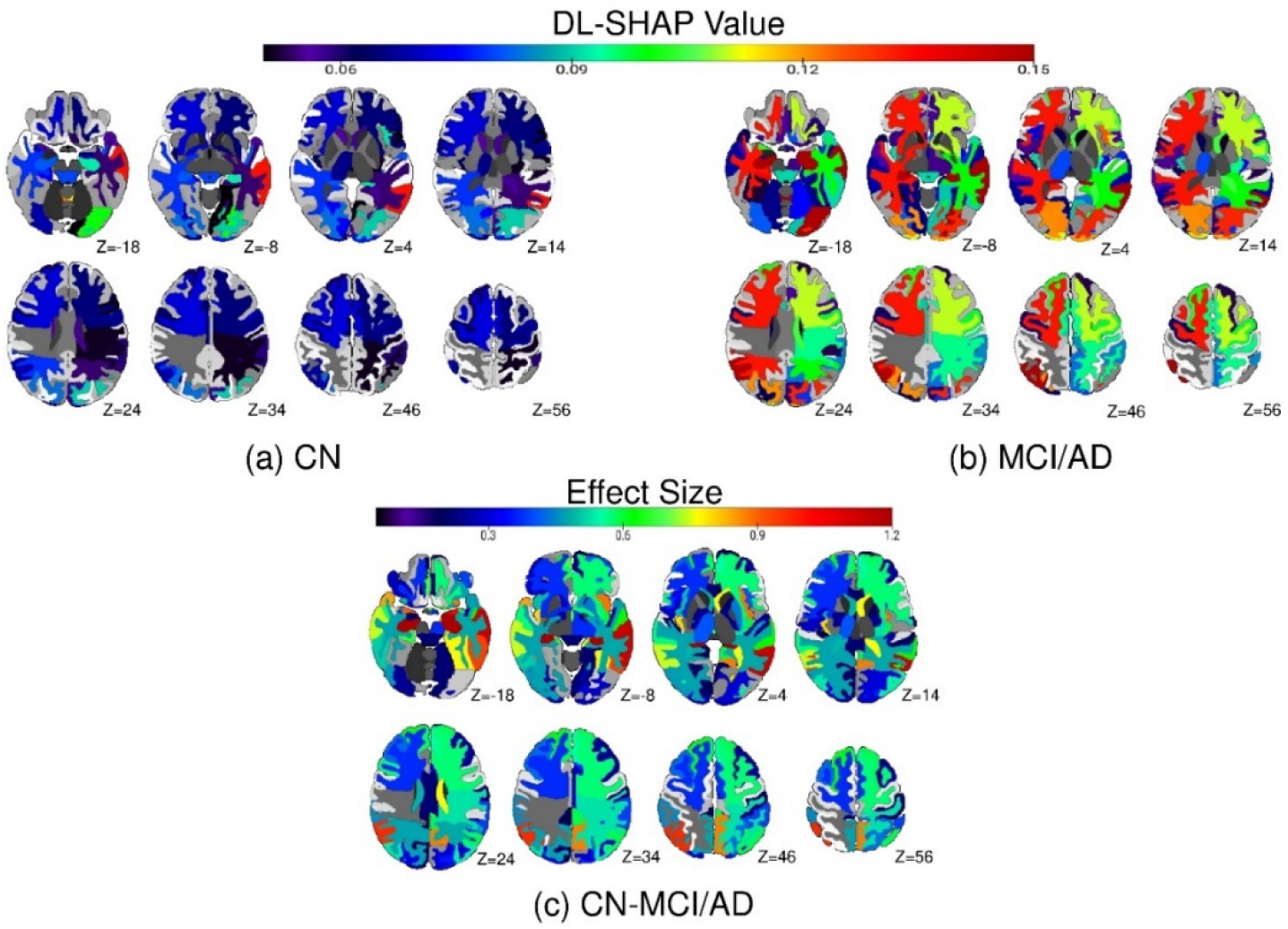
DL-SHAP identified significant brain regions for predicting global cognition (MMSE) using the **Knight-ADRC** experimental data across the **(a)** CN, and **(b)** MCI/ADgroups. The results are averaged over all participants. The Cohen’s d effect size differences in regional DL-SHAP values are shown between groups: **(c)** CN and MCI/AD. z shows the brain slice position.

Furthermore, the differences in feature importance, based on the DL-SHAP values of the input ROIs, between control and patient groups were compared using Cohen’s d method for each data cohort. Like, **Figure 4(d)** and **(e)** reflect the Cohen’s d differences between CN and patient groups (MCI and AD) of ADNI data. As seen in **Figure 4(d)**, the prominent regions with notable effect sizes based on the difference between CN and MCI were the left inferior lateral ventricle, left hippocampus, left amygdala, right hippocampus, and left inferior temporal gyrus, while the dominant regions for the difference between CN and AD, as observed in **Figure 4(e)**, were the left hippocampus, left amygdala, right hippocampus, left middle temporal gyrus, and left inferior lateral ventricle. We noticed that the differences between CN and MCI were smaller. In contrast, the CN vs. AD differences were more pronounced. Additionally, the Cohen’s d difference between CN and MCI/AD of Knight-ADRC is illustrated in **Figure 5(c)**, and the regions with significant effect sizes for this difference were the left hippocampus, right hippocampus, left amygdala, right inferior lateral ventricle, and left middle temporal gyrus. A large brain regional atrophy pattern can be witnessed in this group difference.

### AD’s clinical severity results

The relevance of individualized multivariate regional DL-SHAP features with clinical severity of AD was investigated by computing the Spearman’s correlations of these features with individuals’ CDR-SB score for each cohort respectively, and only the statistically significant correlations (p-value < 0.05) were considered. As demonstrated in **Figure 6(a)**, the brain regions with notable positive correlations with CDR-SB in ADNI were the left precuneus, third ventricle, left middle temporal gyrus, right inferior lateral ventricle, and left precentral gyrus medial segment, whereas the prominent regions in Knight-ADRC, as shown in **Figure 6(b)**, were the left precuneus, right angular gyrus, left amygdala, left precentral gyrus medial segment, and right entorhinal area.

**Figure 6:**
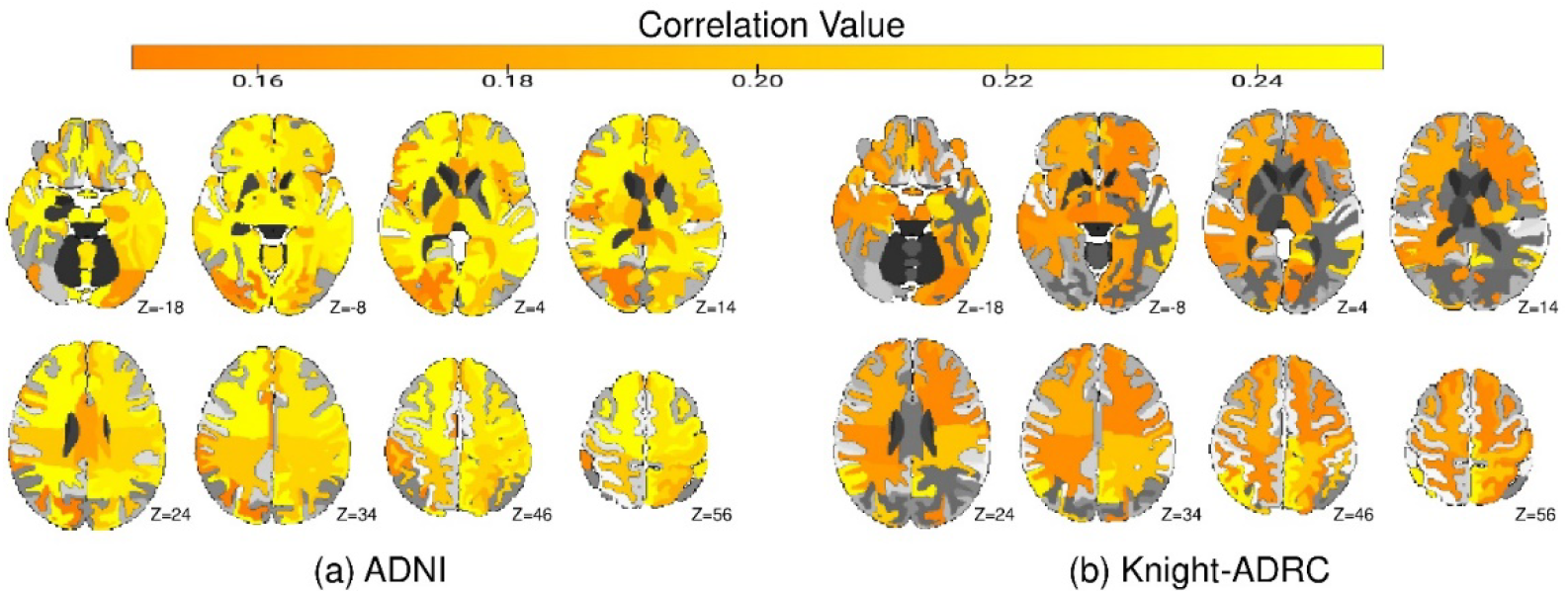
Spearman’s correlations between the individualized DL-SHAP regional features and CDR-SB scores across the AD continuum (CN, MCI, AD) on the: **(a)** ADNI data and **(b)** Knight-ADRC data. The color bar highlights the correlation values and z shows the brain slice position.

## Discussion

This study systematically investigated multivariate regional WBN-cognitive-clinical severity relationships in the AD continuum using explainable ML/AI approaches. We compared a set of ML/AI models for predicting global cognition in the AD continuum using regional neurodegenerative markers from two large-scale, diverse cohorts. Our DL model outperformed conventional ML (LR, RR, and SVR) models in predicting global cognition on both semi-simulated and experimental data. The optimal predictive DL model was then combined with SHAP to form DL-SHAP, which successfully recovered ground-truth multivariate regional WBN-cognitive patterns in the simulated data. The DL-SHAP method further identified multivariate regional WBN-cognitive mapping patterns across both cognitively normal and MCI/AD patient groups. Furthermore, the associations between individualized DL-SHAP-based multivariate regional features and CDR-SB scores suggest their clinical relevance.

The DL model achieved a higher correlation between the actual and predicted cognition scores as compared to the conventional ML (LR, RR, and SVR) models. This is broadly in line with the DL model outperforming in larger-sample settings and high-dimensional data features [63]. Our DL-SHAP method identified different brain regions to be significant for cognition prediction in CN and patient groups of each data cohort, respectively. Our results indicated that many of these regions overlapped across the diagnostic groups within a cohort. For instance, in ADNI, the left middle temporal gyrus, left inferior lateral ventricle, and right cerebellum white matter were among the top five significant regions in all categories. However, the magnitude and order of significance varied across groups, like the significance of the right cerebellum white matter dropped from MCI to AD. A similar pattern was also observed in Knight-ADRC data, where although the left middle temporal gyrus, left inferior lateral ventricle, right cerebellum white matter, and left cerebellum white matter appeared to be in the top five dominant regions for both controls and patients, their importance changed from one category to another. Overall, these findings suggest that as the disease progresses, it alters the brain structure in a way that the associations of brain regions with cognition change. Additionally, in both data cohorts, the same diagnostic categories share several prominent regions, like the left middle temporal gyrus, right cerebellum white matter, and left inferior lateral ventricle were in the top five significant regions for the CN category of both ADNI and Knight-ADRC. Similarly, the AD group of ADNI and the MCI/AD group of Knight-ADRC had all the same top five prominent regions. This indicates the effectiveness of our method to capture the same patterns irrespective of the data diversity, hence proving the generalizability and validity of our technique.

The key cognition-related brain regional hubs identified by DL-SHAP in the AD continuum were notably consistent with the existing literature. For instance, [64] showed the left middle temporal gyrus to be highly correlated with cognitive decline in AD, [65] linked atrophy in the hippocampal region with cognitive impairment, and [66] reported the association of the cerebellum white matter with cognition. Similarly, our model demonstrated these regions to be significant for global cognition in the AD continuum, along with capturing some novel dominant regions, based on their DL-SHAP scores. Additionally, the brain regions exhibiting relationships with clinical severity of AD based on our DL-SHAP individualized features also align with prior studies. For example, the alterations in precuneus and amygdala regions were shown to impact cognition [67, 68], and our analysis also illustrated them to be highly correlated with the CDR-SB scores. These results not only proved the validity of our method by portraying the overlap with previous works but also provided new insights into the identified multivariate novel brain regions hierarchically impacting global cognitive and clinical or disease severity patterns in the AD continuum.

It is worth noting that there were significant overlaps and some variations in the related results between cohorts. While the DL model achieved a very high correlation of 0.981 between the actual and predicted cognition on ADNI data, the correlation (0.892) was slightly lower on Knight-ADRC data. This relatively lower performance on Knight-ADRC data could stem from a higher data imbalance between the CN and patient groups in this cohort, along with data having slightly different age range when compared to ADNI. Moreover, on ADNI data, the Cohen’s d analysis revealed a greater regional atrophy difference between CN and AD groups as compared to CN and MCI groups, indicating that with the progression of disease, larger multivariate neurodegeneration patterns emerge in the brain. Since the Knight-ADRC data lacks distinct MCI and AD categories, making such a comparison is not possible; however, a promising difference was observed between the CN and MCI/AD categories of this cohort. Furthermore, the clinical severity-based prominent brain regions across cohorts were overlapping, such as the left precuneus and left precentral gyrus medial segment, demonstrating our method’s ability to capture similar patterns across cohorts, aiding in proving the validity of our findings and methods. We also observed that the top correlations between the DL-SHAP features and CDR-SB scores on ADNI cohort were higher than the correlations on Knight-ADRC cohort, and this could be due to the larger number of dementia patients in ADNI and thus higher disease or clinical severity patterns in ADNI.

In conclusion, our interpretable deep learning framework, DL-SHAP, showcased high performance in predicting global cognition in the AD continuum using the neurodegenerative markers extracted from a large sMRI dataset belonging to two diverse data cohorts and identified the hierarchy of multivariate WBN regional hubs significantly contributed to global cognitive impairment processes. We also emphasized the relevance of our DL-SHAP features to the clinical or disease severity patterns of MCI/AD relative to controls. Uncovering complex multivariate WBN-cognition maps, where highly significant brain regions likely represent early disease impact and should be prioritized for targeted interventions, can aid in the development of personalized strategies for early detection, delay, or mitigation of cognitive decline in MCI/AD. In the future, applications of DL-SHAP to other neuroimaging modalities or a combination of modalities may capture multiple aspects of brain functions and structures, offering a more comprehensive understanding of AD mechanisms. This multimodal, multiregional approach may reveal novel biomarkers and refine therapeutic strategies, ultimately contributing to more effective management of AD and related neurodegenerative disorders. Our work lays a foundation for future research in WB-cognitive-clinical severity mapping, highlighting DL-SHAP’s potential in advancing both scientific understanding and clinical approaches to neurodegenerative diseases like AD.

## Data Availability

Original ADNI (https://adni.loni.usc.edu/) and Knight-ADRC (https://knightadrc.wustl.edu/) data are publicly available. The generated results will be shared upon reasonable request following applicable human subjects’ data transfer procedures.

## Code Availability

Our code is available at https://github.com/ganchand/DL-SHAP. For harmonization we have used the code located at https://github.com/ganchand/AJP_Codes. The MUSE code is located at https://github.com/CBICA/MUSE.

