## Supplementary Materials for "Multivariate whole brain neurodegenerative-cognitive-clinical severity mapping in the Alzheimer’s disease continuum using explainable AI"

**Supplementary information**

**S1: Semi-simulated data generation**

The simulated data was generated using the sMRIs of cognitively normal (CN) participants belonging to the ADNI-2 and Knight-ADRC cohorts (n=1,107). From these sMR-images, the neurodegeneration volumetric markers corresponding to 145 regions of interest (ROIs) were extracted employing the MUSE tool [1], and then harmonized based on a regression-based harmonization technique [2-4] to eliminate the covariates effects. After that, the neurodegenerative data was normalized for each participant by dividing each ROI value by the sum of all ROIs for eradicating the scale differences and making the data more suitable for AI methods. To generate the simulated data, the top ten ROIs portraying high Spearman correlations with the cognitive score (MMSE) were selected for perturbation. Those ROIs were the left middle temporal gyrus, left middle frontal gyrus, left central operculum, left medial frontal cortex, left supramarginal gyrus, right middle frontal gyrus, right superior temporal gyrus, left anterior insula, right middle temporal gyrus, and right parietal operculum. We then generated simulated data by randomly reducing the volume of the selected ROIs by a factor between 40% and 60% and adjusting the respective cognitive scores by a factor between 1% and 30%, based on the assumption that brain volume and cognitive function (mini-mental state examination (MMSE)) are positively correlated [5-7]. These perturbations were applied to 45% of the participants. We perturbed the neurodegeneration markers and their respective cognitive scores such that the correlation of the ten selected ROIs exhibited a strong and significant correlation with the cognitive measure, while the other ROIs illustrate weak correlations.

**S2: DL model hyperparameter tuning**

To optimize the hyperparameters of the DL model, we conducted a systematic search across a predefined range of candidate values. The selection of optimal hyperparameters was guided by model performance on the MMSE prediction task using the semi-simulated dataset. Performance was assessed using two primary metrics: (1) Spearman’s rank correlation coefficient between the actual and predicted MMSE scores, and (2) Test loss, calculated as the Mean Absolute Error (MAE). To determine the statistical significance of the observed Spearman correlation, we computed the associated p-value, thereby confirming whether the correlation was unlikely to have occurred by chance.

Each hyperparameter was tuned individually by varying its values while keeping all other hyperparameters fixed at their default settings. For each configuration, the model was trained and evaluated across five independent runs, with each run employing a 10-fold cross-validation (CV) training and evaluation procedure. This setup ensures that the model’s performance is not biased by a single data split or initialization. The final reported results represent the average performance across the five runs, thereby enhancing robustness and reducing the impact of randomness or noise in training and evaluation.

The default values of the hyperparameters were as follows: batch size=64, optimizer=ADAM, training epoch=1000, model layers=5 (units: 40, 30, 20, 10, 1), learning rate=0.05, and loss= MAE. For each CV iteration, 9 folds (90% of the data) were used for training and the remaining fold (10%) was reserved exclusively for testing, ensuring a strict separation between training and test data. Within each training fold, we used the validation_split=0.05 parameter in the model.fit() function, in combination with monitor='val_loss' in the EarlyStopping callback. This configuration applied the validation split only to the training data (X_train and y_train) within that fold. The early stopping monitored the loss on this internal validation set and halted training if no improvement was observed for 100 consecutive epochs and restores the model’s best weights. Importantly, the test fold was never used for training or validation during this process. Therefore, the early stopping mechanism was entirely nested within the training data of each CV fold. This ensures that the validation data used for early stopping was fully independent of the test fold, preventing data leakage and avoiding any inflation of the reported test performance. Final evaluation metrics, such as Spearman correlation, were calculated strictly on the held-out test fold after training was complete. Due to dataset size constraints, we limited the internal validation split to 5% in order to retain a sufficiently large training set per fold and ensure model stability.

**Table S2.1** presents the performance of the DL model for MMSE prediction across different learning rates. The results indicate that a learning rate of 0.05 yields the highest Spearman correlation between the actual and predicted MMSE scores. Additionally, the lowest test loss (MAE) is also achieved at this learning rate. These findings suggest that 0.05 is the optimal learning rate, and accordingly, this value was used in all analyses reported in the main manuscript.

**Table S2.2** shows the performance of our DL model for MMSE prediction across various batch sizes. The results indicate that the model achieved the highest Spearman correlation and the lowest test loss (MAE) when the batch size was set to 64. Based on this optimal performance, a batch size of 64 was selected for use in the final model configuration, and all results reported in the main manuscript are based on this setting.

**Table S2.3** demonstrates the performance of our DL model for MMSE prediction using different numbers of layers. The results indicate that the model configuration with five layers, comprising units [40, 30, 20, 10, 1], achieved the highest Spearman correlation and the lowest test loss (MAE). Based on this optimal performance, this architecture was adopted as the final model configuration for reporting the results presented in the main manuscript.

| **Learning Rate** | **Spearman Correlation** | **p-value** | **Test Loss** |
| --- | --- | --- | --- |
| 0.1 | 0.901 | < 2.2E-308 | 1.086 |
| **0.05** | **0.940** | **< 2.2E-308** | **0.872** |
| 0.01 | 0.889 | < 2.2E-308 | 1.686 |
| 0.0045 | 0.897 | < 2.2E-308 | 1.549 |
| 0.001 | 0.841 | 6.67E-298 | 3.500 |

**Table S2.1:** The performance of the DL model with different **learning rates** for MMSE prediction. Spearman’s correlation between the actual and predicted MMSE scores is reported for each learning rate-wise configuration. The optimal performing configuration is shown in bold.

| **Batch Size** | **Spearman Correlation** | **p-value** | **Test Loss** |
| --- | --- | --- | --- |
| 128 | 0.864 | < 2.2e-308 | 4.185 |
| **64** | **0.940** | **< 2.2e-308** | **0.872** |
| 32 | 0.904 | < 2.2e-308 | 1.593 |
| 25 | 0.906 | < 2.2e-308 | 1.690 |

**Table S2.2:** The performance of the DL model with different **batch sizes** for MMSE prediction. Spearman’s correlation between the actual and predicted MMSE scores is reported for each batch size-wise configuration. The optimal performing configuration is shown in bold.

| **# of Layers [Units]** | **Spearman Correlation** | **p-value** | **Test Loss** |
| --- | --- | --- | --- |
| 6 [50, 40, 30, 20, 10, 1] | 0.896 | < 2.2e-308 | 1.816 |
| **5 [40, 30, 20, 10, 1]** | **0.940** | **< 2.2e-308** | **0.872** |
| 4 [30, 20, 10, 1] | 0.907 | < 2.2e-308 | 1.668 |
| 3 [20, 10, 1] | 0.906 | < 2.2e-308 | 1.785 |

**Table S2.3:** The performance of the DL model with different **number of layers** for MMSE prediction. Spearman’s correlation between the actual and predicted MMSE scores is reported for each number of layers-wise configuration. The optimal performing configuration is shown in bold.

**S3: Performance comparison of feature importance techniques**

The performance of three feature importance techniques were compared to determine which technique can best capture the multivariate significant brain regions associated with global cognition (MMSE) prediction. Those interpretability techniques are Shapley Additive Explanations (SHAP) [8], Local Interpretable Model-Agnostic Explanations (LIME) [9, 10], and Layer-wise Relevance Propagation (LRP) [11].

SHAP leverages Shapley values from cooperative game theory to quantify feature importance in predictive models. By assessing the marginal contribution of each feature across all possible feature combinations, SHAP provides a consistent and theoretically justified measure of feature influence on model output.

LIME interprets individual predictions by fitting an interpretable surrogate model, typically linear, to locally approximate the original complex model. It generates perturbed samples around the target instance and trains the surrogate to replicate the model’s local behavior. The explanation model for feature x is obtained by solving:

$$explanation \left( x \right)=\arg{min}_{g\in G} L\left( f, g,\pi_{x} \right)+ Ω(g)$$

where f is the original model (e.g., DL), g is a simple explanation model from the family G, L is the loss function measuring the fidelity between f and g in the local neighborhood defined by π_x_, and Ω(g) penalizes the complexity of g.

LRP is a technique for interpreting deep neural networks by backpropagating the output prediction through the network layers to the input features, attributing relevance scores that reflect each feature’s contribution. The relevance $R_{i}$ for input feature $x_{i}$ is computed using the formula:

$$R_{i} = \sum_{j} \frac{x_{i}w_{ij}}{\sum_{k} x_{k}w_{kj+ \epsilon.sign(\sum_{k} x_{k}w_{kj})}} . R_{j}$$

where $R_{j}$ is the relevance score of neuron j in the next layer, w_ij_ are the weights, and ε is a small constant added for numerical stability. The method follows the relevance conservation principle, ensuring that the total relevance is preserved as it is distributed backward through the network.

To validate the performance of these feature attribution methods, we conducted experiments using the semi-simulated data for MMSE (Mini-Mental State Examination) prediction with our deep learning (DL) model. Specifically, we integrated the DL model with each of the respective explainability techniques to form DL-LIME, DL-LRP, and DL-SHAP models. Since the ground-truth perturbed brain regions in the semi-simulated data are known and represent pseudo-patient patterns, we used them as a benchmark to assess how accurately each method could identify these regions to be significant for the MMSE predictions. **Table S3.1** presents the ground-truth perturbed regions and indicates whether these were ranked among the top 10 most significant regions identified by each interpretability method. As shown, DL-SHAP successfully captured **100%** of the ground-truth regions in its top 10 features, compared to **80%** by DL-LIME and **70%** by DL-LRP. These results demonstrate that SHAP provides the most accurate identification of the significant brain regions relevant to MMSE prediction in our experimental setting, therefore we chose SHAP technique in this work. In addition, **Figure S3.1** illustrates the top 10 significant brain regions identified by **(a)** DL-LRP, **(b)** DL-LIME, and **(c)** DL-SHAP. Notably, all the regions highlighted by DL-SHAP overlap with the known ground-truth perturbed regions. Furthermore, we implemented LRP, LIME, and SHAP techniques in Python. For SHAP, we used the Explainer interface. For LIME, we utilized the LimeTabularExplainer interface with the feature_selection='lasso_path', mode='regression', num_features=22, and num_samples=2000. For LRP, the innvestigate library with lrp.epsilon, where epsilon=1e-7, configuration is employed. For each technique, we ran the model 5 times and took the averaged absolute values for deciding the feature importance respectively.

| **Perturb Regions** | **Top 10 DL-LRP Regions** | **Top 10 DL-LIME Regions** | **Top 10 DL-SHAP Regions** |
| --- | --- | --- | --- |
| Left Middle Temporal Gyrus | ✓ | ✓ | ✓ |
| Left Middle Frontal Gyrus | ✓ | ✓ | ✓ |
| Left Central Operculum | ✓ | ✓ | ✓ |
| Left Medial Frontal Cortex | ✖ | ✓ | ✓ |
| Left Supramarginal Gyrus | ✓ | ✓ | ✓ |
| Right Middle Frontal Gyrus | ✓ | ✖ | ✓ |
| Right Superior Temporal Gyrus | ✖ | ✓ | ✓ |
| Left Anterior Insula | ✓ | ✓ | ✓ |
| Right Middle Temporal Gyrus | ✓ | ✓ | ✓ |
| Right Parietal Operculum | ✖ | ✖ | ✓ |

**Table S3.1**: The list of perturbed brain regions representing the pseudo-patient patterns in semi-simulated data, and the indication of those regions being in the top 10 significant brain regions by DL-LRP, DL-LIME, and DL-SHAP respectively.

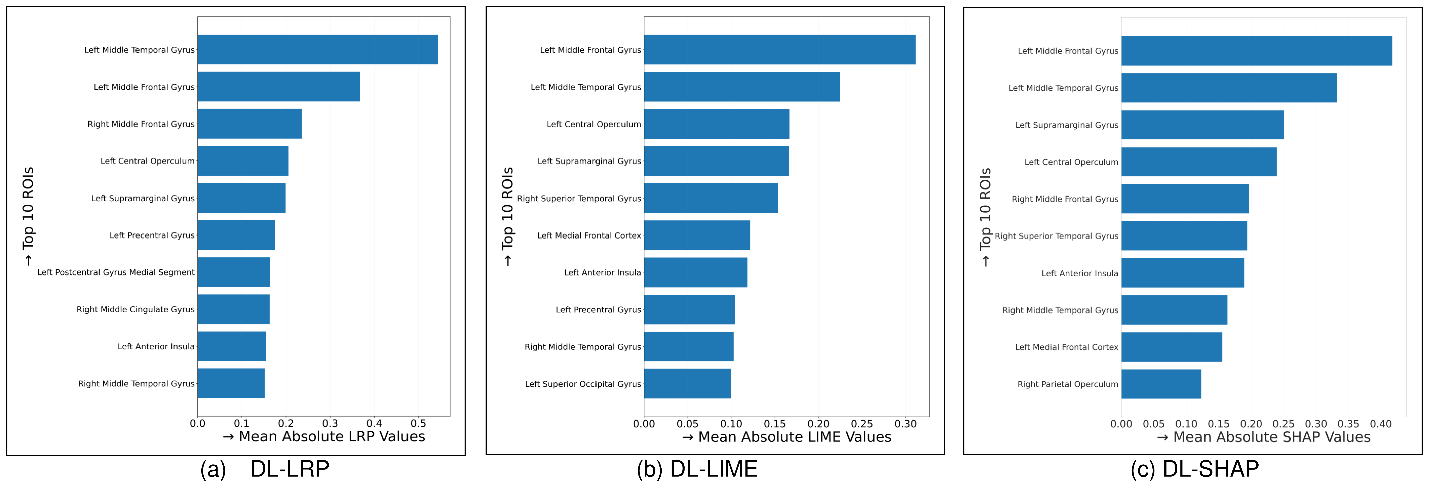

**Figure S3.1:** The top 10 significant brain regions captured by **(a)** DL-LRP, **(b)** DL**-**LIME, and **(c)** DL-SHAP, respectively, for MMSE prediction using the semi-simulated data.
